# All text: A Novel Scoring System for Precise Severity Quantification in Severe Fever with Thrombocytopenia Syndrome: Development and Application Based on Dynamic Clinical Data

**DOI:** 10.64898/2026.02.17.26346452

**Authors:** Yingchun Sun, Zhiyu Pan, Jing Sun, Yihui Sun, Wenjie Wang, Manman Liang, Aiping Zhang, Qiongle Wu, Haoyu Sheng, Jianghua Yang

**Author notes:** Yingchun Sun, Zhiyu Pan, Jing Sun contributed equally to this work. **Correspondence:** Jiang-Hua Yang.

## Abstract

**Background:** Severe Fever with Thrombocytopenia Syndrome (SFTS) is an acute infectious disease with high mortality. This study aimed to develop a quantitative scoring system for grading SFTS severity using dynamic clinical data.

**Methods:** A retrospective study included 547 confirmed SFTS patients from two hospitals. Clinical data were collected over a 14-day course (divided into four phases). Patients were grouped into survivors (n=451) and non-survivors (n=96). Statistical analyses, including Kaplan-Meier curves and log-rank tests, were performed. An external validation cohort of 44 new patients was used to validate the scoring system via C-statistic, calibration curves, and decision curve analysis (DCA).

**Results:** Of 547 patients, 96 (17.55%) were non-survivors. Multivariate logistic regression identified six independent prognostic factors across phases: age, platelet (PLT), aspartate aminotransferase (AST), and creatinine (Cr) (days 5-7); age, red blood cell distribution width (RDW), Cr, and lactate dehydrogenase (LDH) (days 8-10); Cr and LDH (days 11-14). A scoring system (0-11 points) was developed, stratifying patients into low (0-3), intermediate (4-7), and high (8-11) risk groups, with adverse outcome rates of 1.04%, 22.92%, and 76.04%, respectively. Kaplan-Meier curves showed significant prognostic differences (log-rank P<0.001). External validation (44 cases) confirmed excellent performance: AUC 0.810-0.952, good calibration (Hosmer-Lemeshow P>0.05), and net clinical benefit (DCA Eavg 0.068-0.098, Emax 0.422-0.559).

**Conclusion:** A dynamic SFTS severity scoring system was developed and validated. Internal and external validation confirmed its reliability and clinical utility, providing a simple, practical tool for timely assessment and early intervention.

## 1. Introduction

Severe Fever with Thrombocytopenia Syndrome (SFTS) is an acute infectious disease caused by *Dabie bandavirus* (DBV), primarily transmitted via tick bites or contact with the blood and bodily fluids of infected individuals, and other routes[2,6,25]. Current research indicates that the case mortality rate of SFTS ranges from 12% to 30% in East Asia[4,19,20,25]. The epidemic area continues to expand to new geographical regions, with both the proportion of severe cases and the mortality rate showing a gradual upward trend[19,20,25]. Thus, it has become a significant regional public health threat in China and several other East Asian countries[1,2,3,4].

The clinical manifestations of SFTS are typically characterized by fever (often persistent high fever, with body temperature frequently ≥38.5°C), thrombocytopenia, leukopenia, and gastrointestinal symptoms (such as nausea, vomiting, abdominal pain, and diarrhea). However, the severity of the disease varies considerably among patients. Some patients only present with mild, self-limiting symptoms, potentially accompanied by non-specific manifestations such as fatigue and muscle aches[3,6,19,25,26]. These cases usually have a short course and favourable prognosis, resolving spontaneously without specific targeted treatment[3,26]. In contrast, others patients may rapidly progress to multi-organ failure (e.g., hepatic failure, renal failure, disseminated intravascular coagulation, shock), requiring urgent comprehensive resuscitative measures including mechanical ventilation, hemodialysis, and anti-shock therapy, yet still having a poor prognosis[6,11,19,25,26]. Therefore, clinical practice requires individualized therapeutic approaches based on the severity of the patient’s condition[22,23,27].

Currently, relevant domestic and international diagnostic and treatment guidelines have proposed preliminary frameworks for assessing the severity of SFTS, broadly classifying patients into mild and severe cases[6,7,26]. However, the relevant criteria primarily rely on the presence or absence of clinical symptoms and thresholds for single laboratory indicators. For example PLT<50 × 10 ⁹ /L and white blood cell (WBC) counts<2 × 10 ⁹ /L are used as reference indicators for determining severe cases[8,9,10,25]. Current research progress shows that there are significant shortcomings in the assessment SFTS severity, most notably the prevalence of cross-sectional study designs[21,22,23,24]. Such studies typically collect laboratory indicators (e.g., PLT, WBC, hepatic and renal function, coagulation parameters) and clinical symptoms only at admission or a single specific time point. Constructing disease assessment models or screening for risk factors based on single-time-point data fails to reflect the dynamic progression of the disease[8,9]. Without timely adjustments to monitoring frequency, this approach may delaying intervention opportunities[21,22,23,24,27].

Therefore, to achieve a dynamic and precise quantitative assessment of disease severity in SFTS patients, this study divided the data of 547 SFTS patients within a 14-day disease course into four phases. By extracting the extreme values of indicators at each phase, the study captured the peak characteristics of disease severity, identified core risk factors for precise quantification, and established a stratified scoring system. Extracting validation with new data provides clinicians with a dynamic, precisely quantifiable, and operationally feasible tools for SFTS disease grading.

## 2. Materials and Methods

### 2.1 Data Collection

A retrospective analysis was conducted on 547 confirmed SFTS patients admitted to the First Affiliated Hospital of Wannan Medical College (Yijishan Hospital) and the First Affiliated Hospital of Nanjing Medical University between May 2020 and June 2024. The patients were divided into a development set (465 cases) and an internal validation set (82 cases) in a ratio of 0.85:0.15. An external validation set included 44 newly diagnosed SFTS patients from the First Affiliated Hospital of Nanjing Medical University between August 2024 and April 2025.

Inclusion criteria: ①Meeting the diagnostic criteria outlined in the Diagnosis and Treatment Protocol for Fever with Thrombocytopenia Syndrome (2020 Edition) (SFTSV nucleic acid positive or specific IgM antibody positive)[7,25];②Age≥18 years.

Exclusion criteria: ①Concurrent infectious diseases (e.g., influenza, sepsis); ② History of severe hepatic or renal insufficiency, hematological disorders, etc.; ③ Clinical data completeness rate<30%.

### 2.2 Staging of the Disease

The course of SFTS was divided into 5 phases based on clinical symptoms and laboratory indicators[5]: Initial phase (days 1-4): Mainly characterized by fever (body temperature≥38.5℃) and mild thrombocytopenia (50-100 × 10⁹/L), without obvious organ dysfunction; Progressive phase (days 5-7): Abnormal liver function (AST>80U/L) or renal function (Cr>100 μmol/L), or mild prolongation of activated partial thromboplastin time (APTT) (>35s), but not meeting the criteria for multiple organ dysfunction; Multiple organ dysfunction (MOD) phase (days 8-10): Associated with 2 or more organ dysfunctions (e.g., acute kidney injury (AKI): Cr>133 μmol/L or urine output<0.5 ml·kg⁻¹·h⁻¹; coagulation dysfunction: APTT>50 s or D-dimer (D-D)>5 mg/L; liver failure: AST>400 U/L or total bilirubin>34 μmol/L); Remission phase (days 11-14): Body temperature returns to normal, and organ function indicators (AST, Cr, APTT) decrease by ≥ 30% compared with the peak; Recovery phase (days 15-20): Not included in the analysis.

### 2.3 Statistical Methods

This study collected a total of 83 variables, including general information, epidemiological data, and laboratory indicators. With prognosis as the dependent variable and extreme values of indicators at various phases as independent variables, univariate logistic regression and LASSO regression were used during development phase to screen for potential prognosis-related variables. The selected variables were then incorporated into multivariate logistic regression analysis, with P<0.05 serving as the inclusion criterion to determine independent risk factors.

Independent risk factors were incorporated into a nomogram, and ROC curves and the area under the ROC curve (AUC) were used to evaluate the predictive accuracy of the model. The overall efficacy of the model was validated through decision curve analysis (DCA).

In the development phase, the K-M curve was used to describe the cumulative incidence of adverse outcomes across risk strata, the log-rank tests were used to compare differences between strata. The C-statistic was calculated to assess the discriminatory power of the scoring system.

In external validation: The C-statistic (95% confidence interval) was calculated to assess discriminatory ability; calibration curves was plotted and calibration was evaluated using the Hosmer-Lemeshow test (P>0.05 indicates good calibration); decision curve analysis (DCA) was constructed to compare the clinical net benefit of different scores.

Statistical analysis was performed using R 4.5.0(rms, glmnet, rmda, pROC, and insert packages) and SPSS Statistics 26.0. Quantitative data are presented as mean± SD or median, with intergroup comparisons performed using t-tests and Mann-Whitney U tests. Categorical data are expressed as frequencies, with intergroup comparisons conducted using χ² tests. Two-tailed p<0.05 was considered statistically significant.

## 3. Results

### 3.1 Baseline Characteristics

Among 547 SFTS patients, 244(44.61%) were male and 303(55.39%) were female, with a median age of 67 years (58-72 years) (P=0.001). The overall mortality rate was 17.55%(96/547). The external validation set (44 patients) showed no significant differences in baseline characteristics compared with the full cohort (P>0.05), ensuring comparability.

Dynamic analysis of clinical and laboratory indicators across four phases of the 14-day disease course revealed statistically significant differences in the distribution of core indicators between the survivor and non-survivor groups at each phase, with the number of differential indicators increasing first and then decreasing with disease progression (Table 1).

**Table 1.**
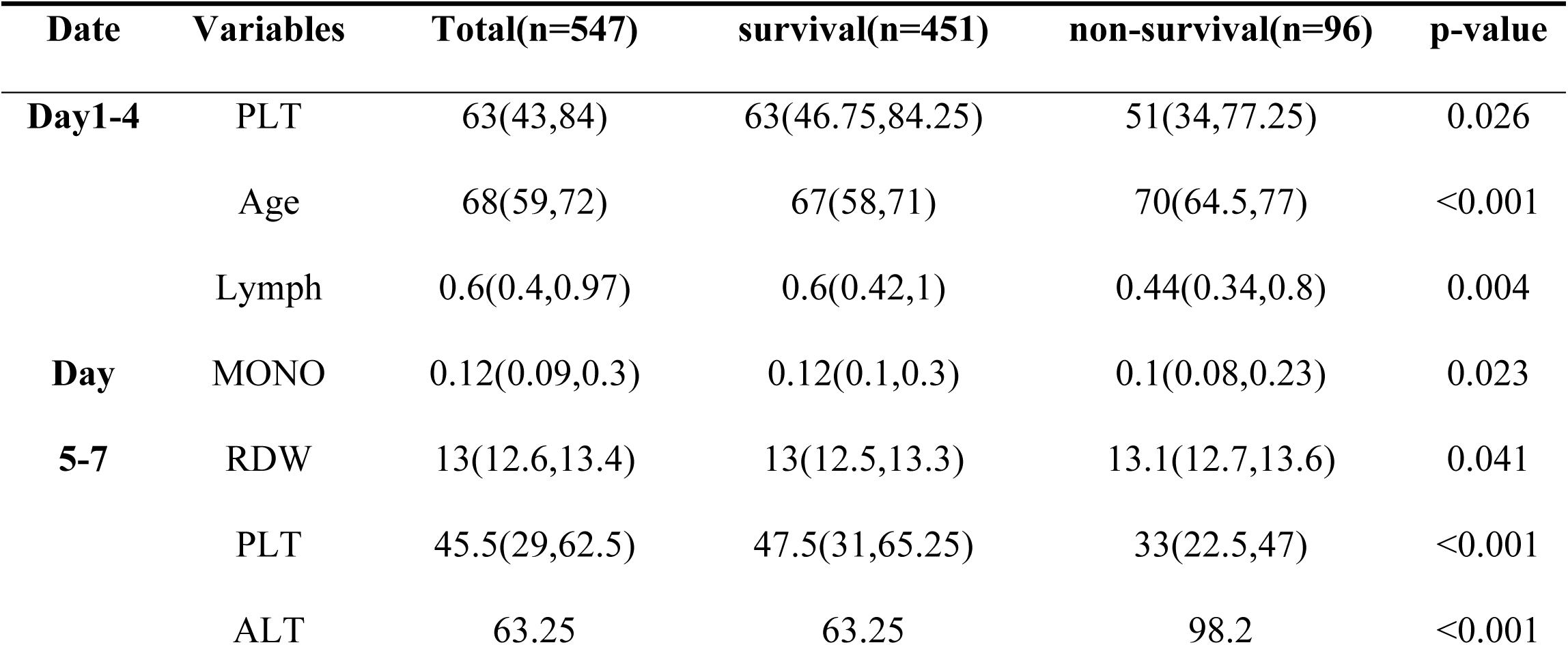
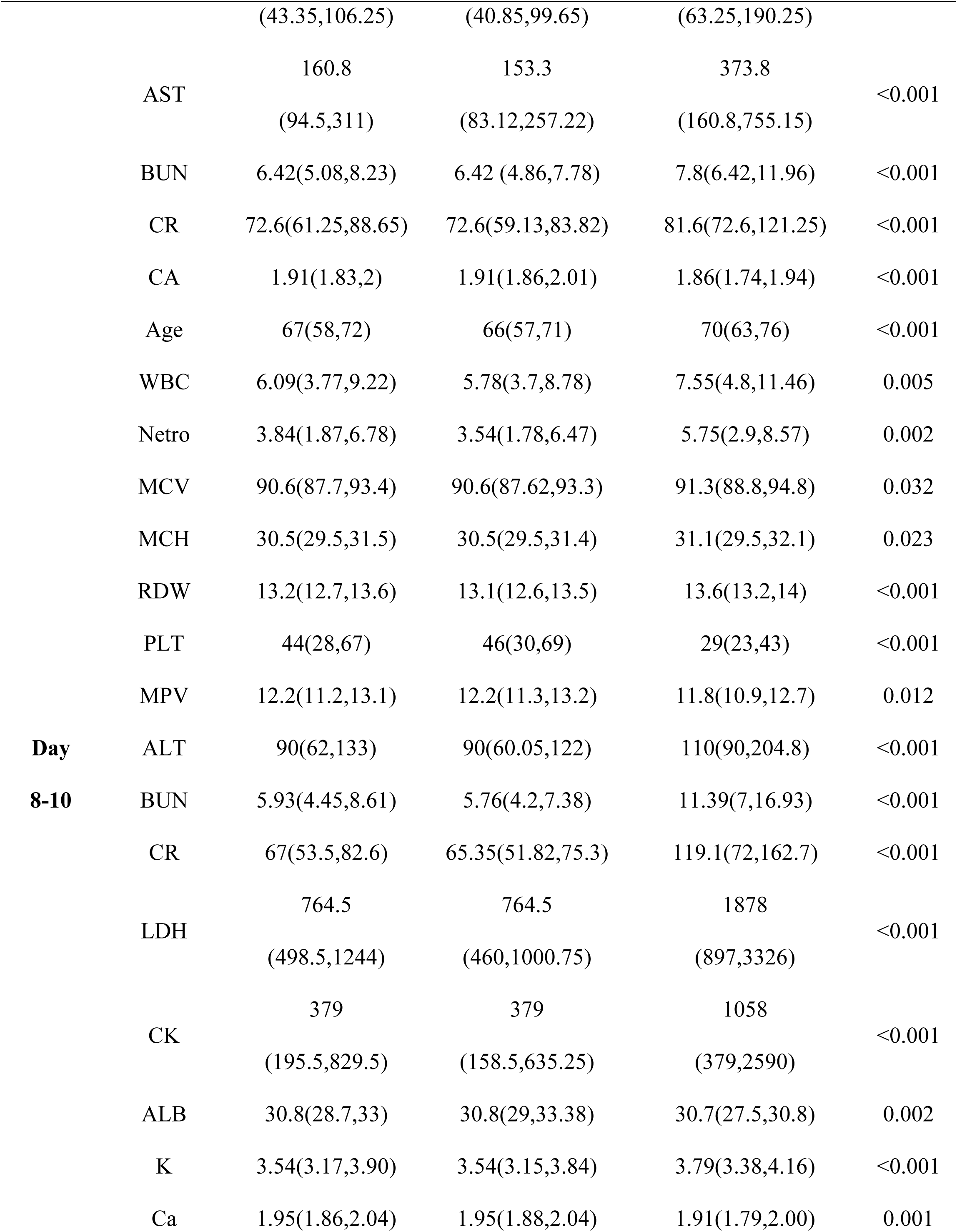
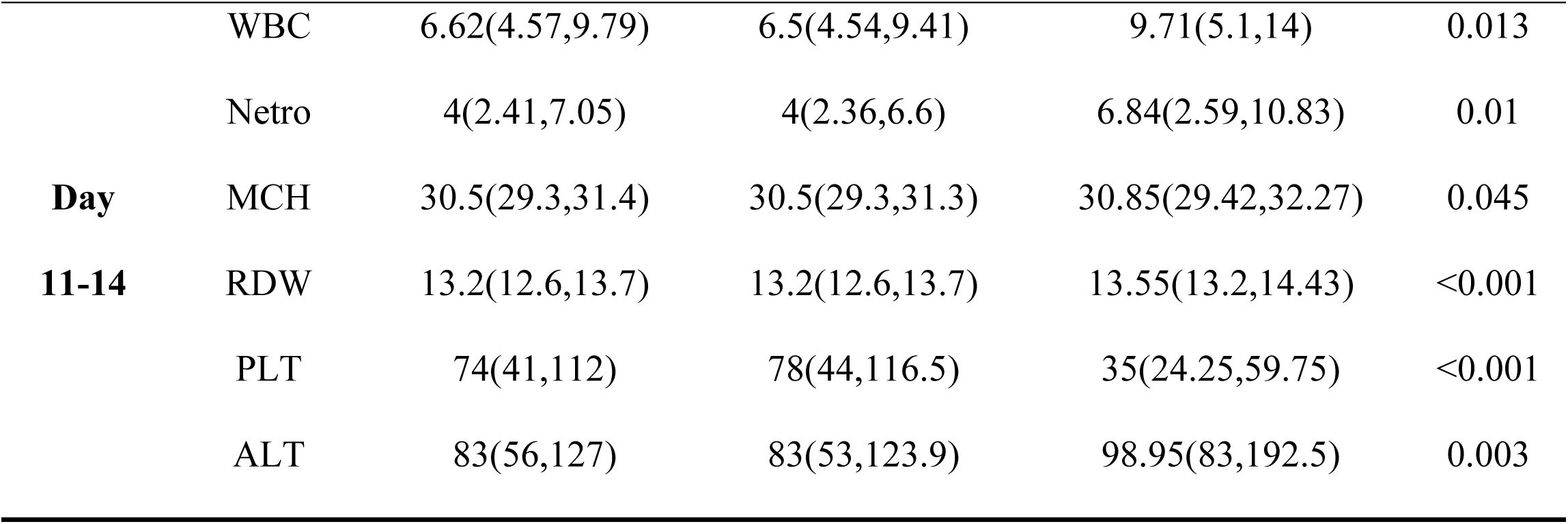
Dynamic analysis of Baseline Characteristics Between Survivor and Non-survivor groups by Disease Phases.

In the initial phase (days 1-4), only platelet count (PLT) showed an intergroup difference (P=0.026), with the non-survivor group having a significantly lower median PLT level.

In the progressive phase (days 5-7), multiple indicators presented intergroup differences (all P<0.05), among which age, PLT, aspartate aminotransferase (AST), and creatinine (Cr) exhibited extremely significant differences (P<0.001); the non-survivor group was characterized by older age, elevated AST and Cr, and reduced PLT.

The multiple organ dysfunction (MOD) phase (days 8-10) had the largest number of differential indicators, covering blood routine parameters, platelet parameters, hepatic and renal function indices, myocardial enzymes, and electrolytes. All indicators showed extremely significant differences (P<0.001) except for mean corpuscular volume (MCV) and mean corpuscular hemoglobin (MCH) (P<0.05), and the non-survivor group presented with elevated WBC, neutrophil (Netro), red blood cell distribution width (RDW), hepatic and renal function indices, myocardial enzymes, and decreased PLT.

In the remission phase (days 11-14), the number of differential indicators decreased, and only WBC, Netro, RDW, PLT, and alanine aminotransferase (ALT) still had intergroup differences (all P<0.05), with the non-survivor group remaining with abnormal core indicators such as decreased PLT and elevated RDW.

### 3.2 Prognosis-Related Factors Across Disease Phases

Univariate analysis revealed significant differences in factors associated with the prognosis of SFTS patient across different disease phases (Table 2):

**Table 2.**
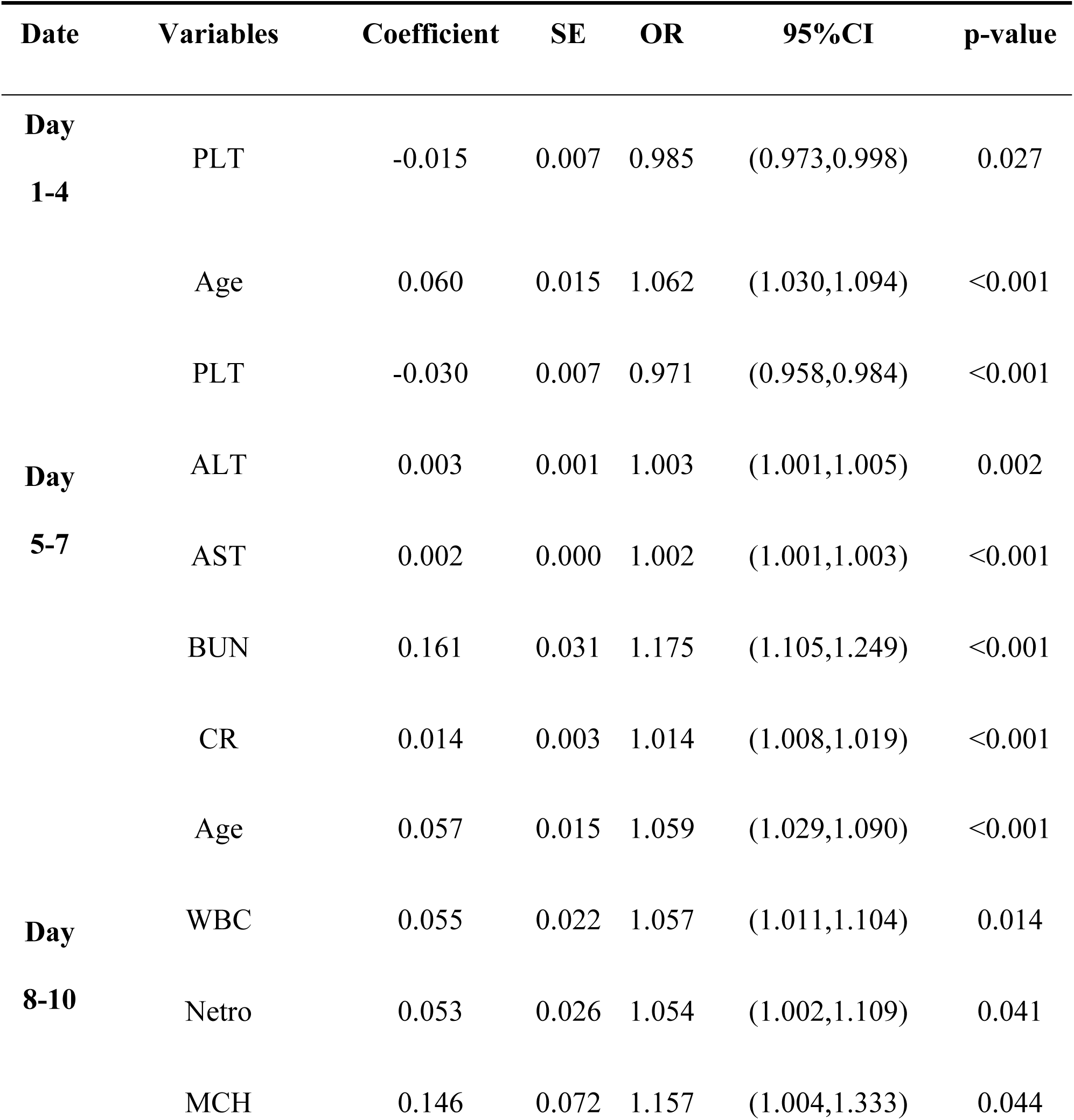
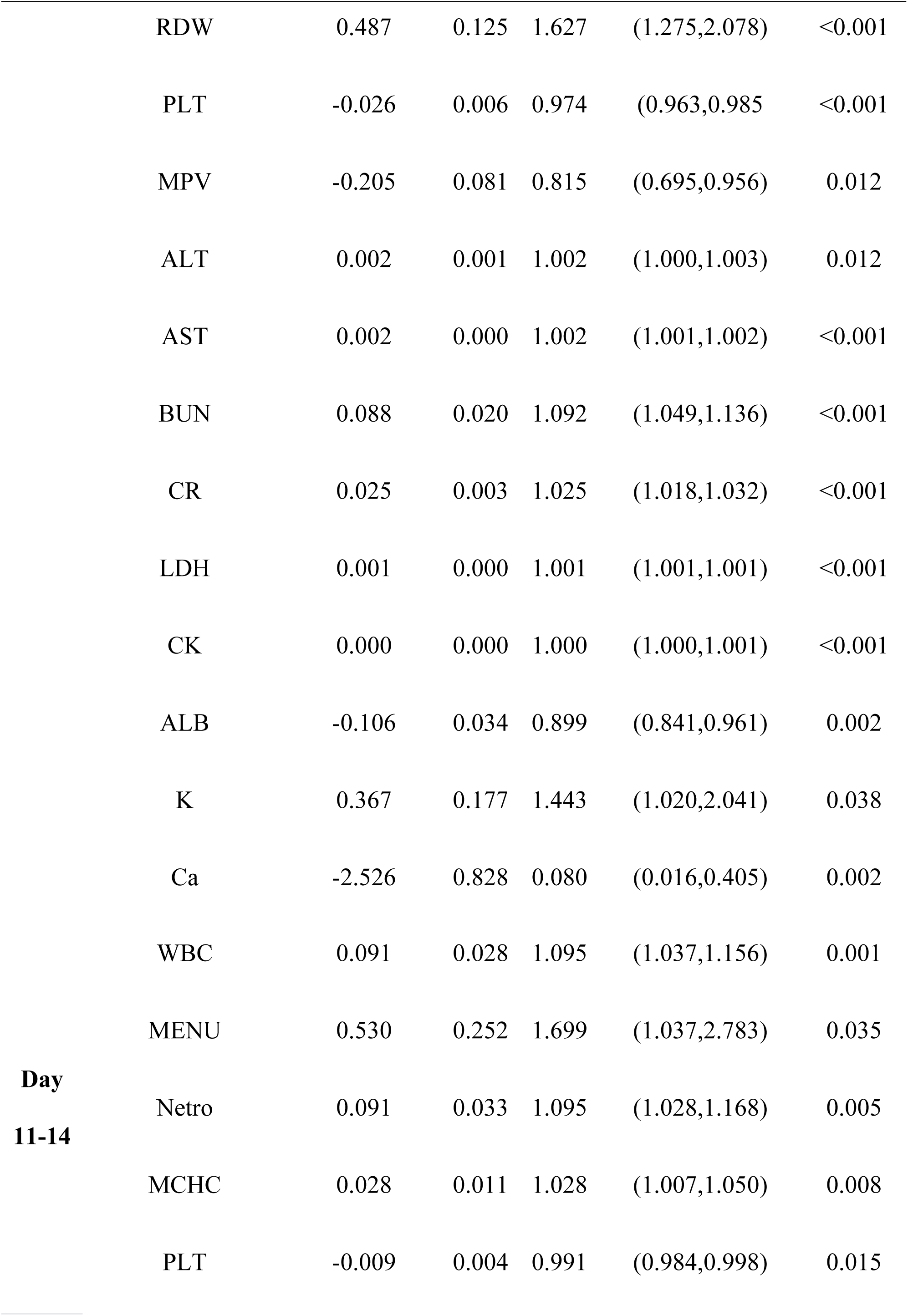
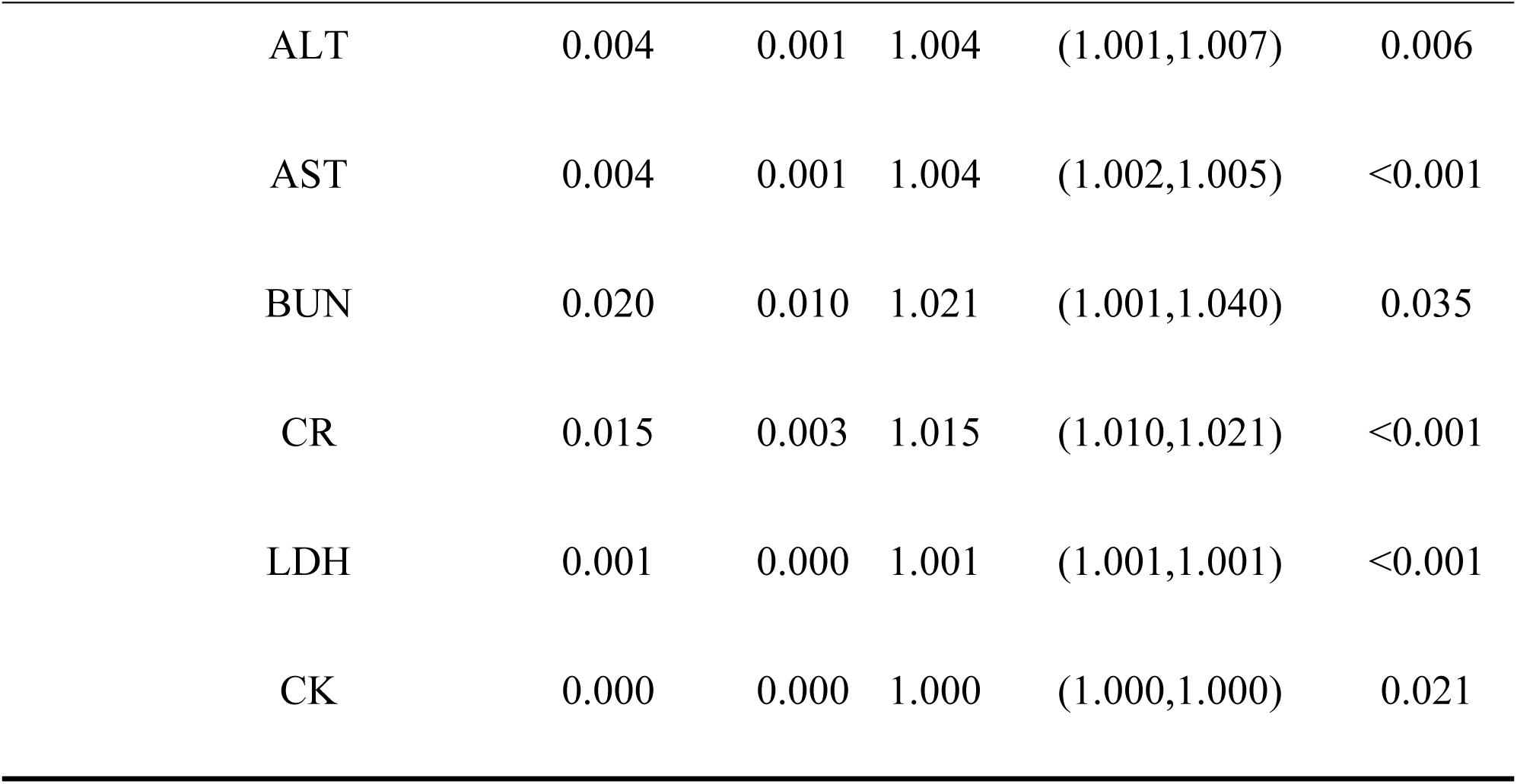
Screening of Prognosis Factors at Different Disease Phases in SFTS Patients.

Days 1-4: Only PLT was associated with prognosis, indicating that decreased PLT levels during this phase are associated with an increased risk of prognosis.

Days 5-7: Six prognostic factors were identified, including age, PLT, alanine aminotransferase (ALT), AST, blood urea nitrogen (BUN), and Cr. Elevated levels of age, ALT, AST, BUN, and Cr, as well as decreased PLT, were all associated with prognosis.

Days 8-10: This phase exhibited the most abundant prognostic factors, totaling 16 items. These included blood count parameters (WBC, neutrophil (Neutro), mean corpuscular hemoglobin (MCH), RDW, PLT, mean platelet volume (MPV)), hepatic and renal function indicator (ALT, AST, BUN, Cr), cardiac enzyme profiles, and metabolic markers (LDH, creatine kinase (CK), albumin (ALB), potassium (K), calcium (Ca)). Among these, RDW, Cr and LDH demonstrated strong predictive value for prognosis.

Days 11-14: Eleven indicators correlated with prognosis, including WBC, Neutro, PLT, AST, Cr and LDH, among others, with particularly significant correlations observed for Cr and LDH.

### 3.3 Independent risk factors for prognosis

Multivariate logistic regression confirmed 6 independent risk factors: age, RDW, PLT, AST, Cr, and LDH. The composition of independent risk factors differed across disease phases (Table 3): Days 1-4: No clear independent risk factors were identified. Days 5-7: Age, PLT, AST and Cr were independent risk factors. Days 8-10: Independent risk factors included age, RDW, Cr, and LDH. Days 11-14: Only Cr and LDH were independent risk factors (Fig 1).

**Figure 1.**
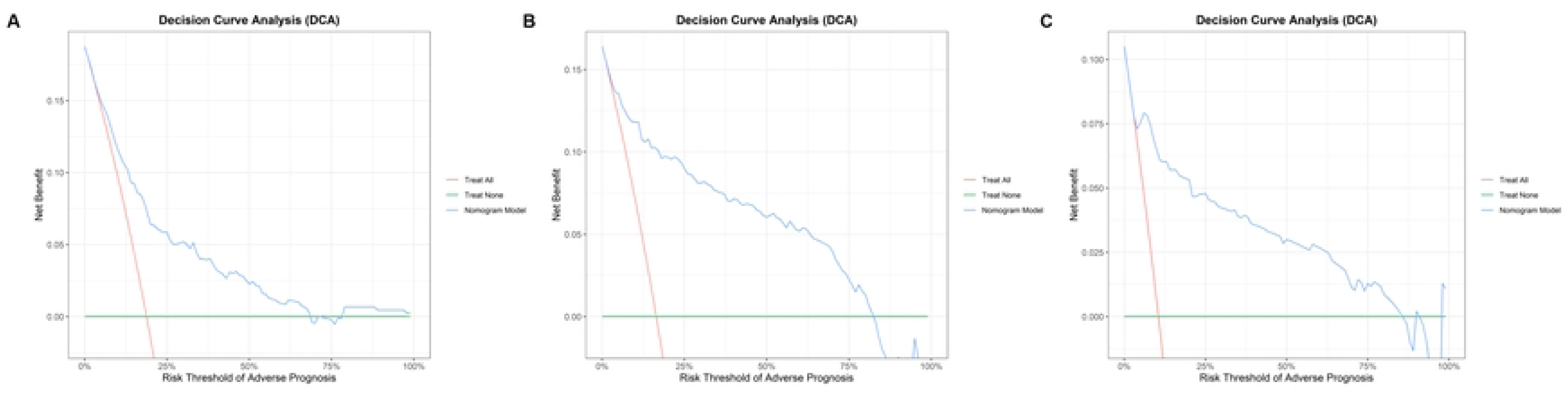
Nomograms. Nomograms for predicting the risk of adverse prognosis in patients with SFTS across different disease phases. (A)Nomogram for predicting during the progressive phase; (B)Nomogram for predicting during the MOD phase; (C)Nomogram for predicting during the remission phase.

**Table 3.** Independent Risk Factors for Prognosis at Different Phases in SFTS Patients.

Patients
| Date | Variables | Coefficient | SE | OR | 95%CI | p-value |
| --- | --- | --- | --- | --- | --- | --- |
| Day |  |  |  |  |  |  |
| 1-4 | - | - | - | - | - | - |
| Day<br>5-7 | Age | 0.0607 | 0.0191 | 1.0625 | (1.0236,1.1028) | 0.0015 |
|  | PLT | -0.0201 | 0.0081 | 0.9801 | (0.9644,0.9961) | 0.0127 |
|  | AST | 0.0009 | 0.0004 | 1.0009 | (1.0001,1.0017) | 0.0155 |
|  | CR | 0.0095 | 0.0030 | 1.0095 | (1.0036,1.0155) | 0.0016 |
| Day<br>8-10 | Age | 0.0504 | 0.0205 | 1.0517 | (1.0105,1.0940) | 0.0140 |
|  | RDW | 0.3496 | 0.1498 | 1.4185 | (1.0573,1.9038) | 0.0196 |
|  | CR | 0.0134 | 0.0034 | 1.0135 | (1.0068,1.0202) | <0.0001 |
|  | LDH | 0.0008 | 0.0002 | 1.0008 | (1.0004,1.0012) | <0.0001 |
| Day<br>11-14 | CR | 0.0090 | 0.0025 | 1.0090 | (1.0041,1.0140) | 0.0003 |
|  | LDH | 0.0004 | 0.0002 | 1.0004 | (1.0000,1.0008) | 0.0496 |

### 3.4 Development of a Severity Scoring System for SFTS Patients

Based on the clinical cut-off values and normal ranges, a stratified scoring rule was established (Table 4): the low-risk tier was assigned 0 points, the medium-risk tier was assigned 1 point, and the high-risk tier was assigned 2 points. For RDW, only low– and medium-risk tiers were assigned (≤14.5% scored 0 points, >14.5% scored 1 point), while all other indicators (PLT, AST, Cr, LDH, and age) were scored across three risk tiers. The total score ranges from 0 to 11 points, categorizing patients into low-risk (0-3 points), intermediate-risk (4-7 points), and high-risk (8-11 points) groups.

**Table 4.** Scoring Rules and Grading Criteria for SFTS Severity Assessment.

| Independent risk factors | 0 points | 1 points | 2 points |
| --- | --- | --- | --- |
| RDW(%) | $\leq 14.5$ | $> 14.5$ | - |
| PLT($\times 10^9/L$) | $\geq 100$ | 50-99 | $< 50$ |
| AST(U/L) | $\leq 40$ | 40-292 | $> 292$ |
| CR($\mu\text{mol/L}$) | $\leq 81$ | 81-103.7 | $> 103.7$ |
| LDH(U/L) | $\leq 250$ | 250-1344 | $> 1344$ |
| Age(years) | $\leq 67$ | 67-73.5 | $> 73.5$ |
| Overall score | 0-3 points<br>(Low risk) | 4-7 points<br>(Medium risk) | 8-11 points<br>(High risk) |

### 3.5 Severity Stratification and Clinical Outcomes

#### 3.5.1 Overall Severity Stratification at 14 days post-admission

Among 547 patients, 99(18.1%) were classified as low risk, 320(58.5%) as intermediate risk, and 128(23.4%) as high risk. The corresponding 14-day adverse outcome rates were 1.04%(1/99), 22.92%(22/320), and 76.04%(73/128), demonstrating a significant risk gradient difference (Table 5). Mortality was extremely low in the low-risk group (0-3 points), with only one death occurring in the 3-point subgroup. The intermediate-risk group (4-7 points) exhibited moderate mortality, with rates increasing slightly across subgroups as scores rose. The high-risk group (8-11 points) showed a markedly elevated mortality rate, with the 11-point subgroup reaching 100% mortality and all 8-10-point subgroups exceeding 50% mortality. This indicates a strong association between higher risk scores and prognosis.

**Table 5.**
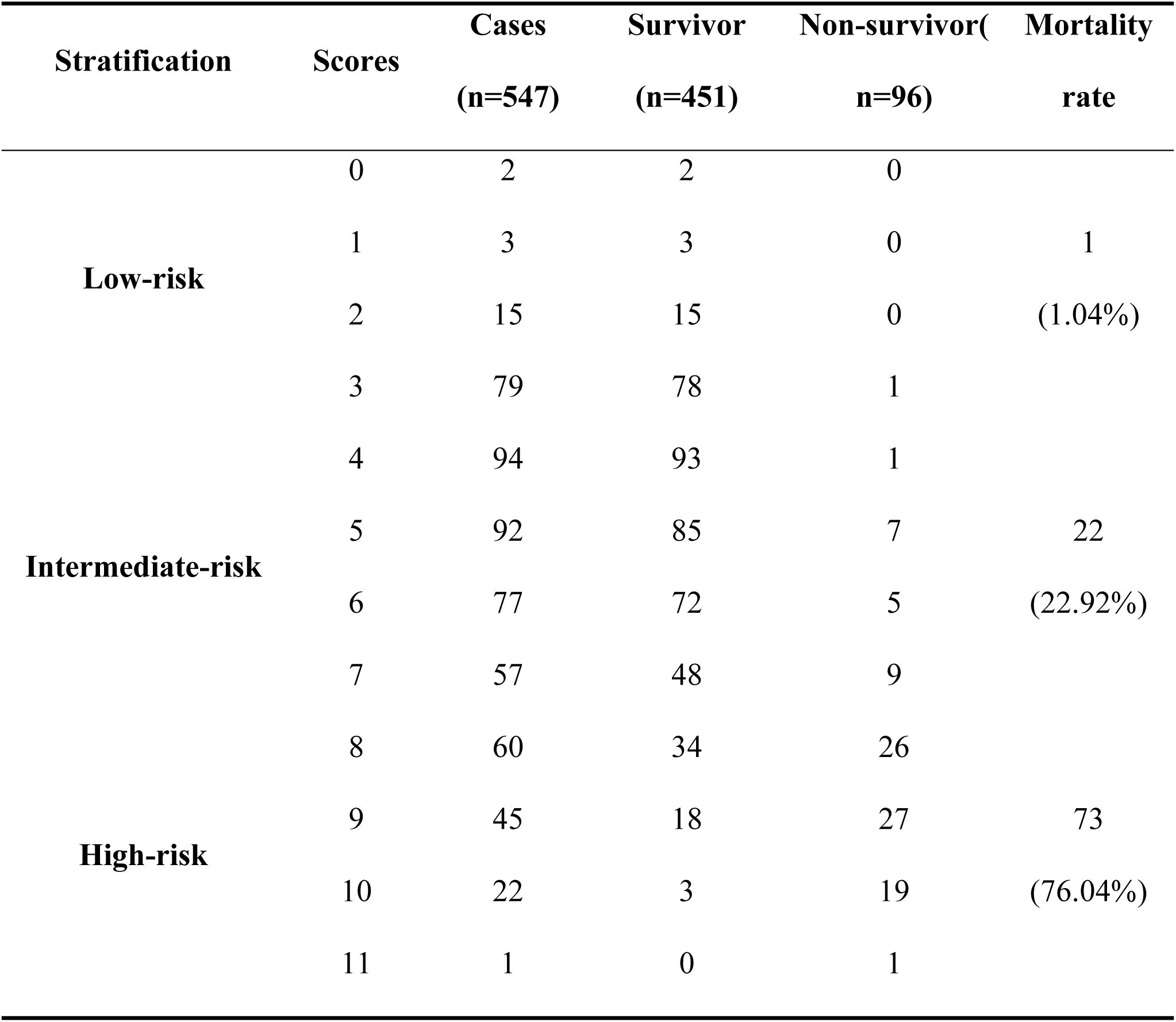
Risk Stratification Scores of SFTS Patients Admitted for 14 Days.

#### 3.5.2 Dynamic Severity Stratification Across Disease Phases

Risk stratification and mortality rates exhibit dynamic correlations across different disease phases (Table 6):

**Table 6.**
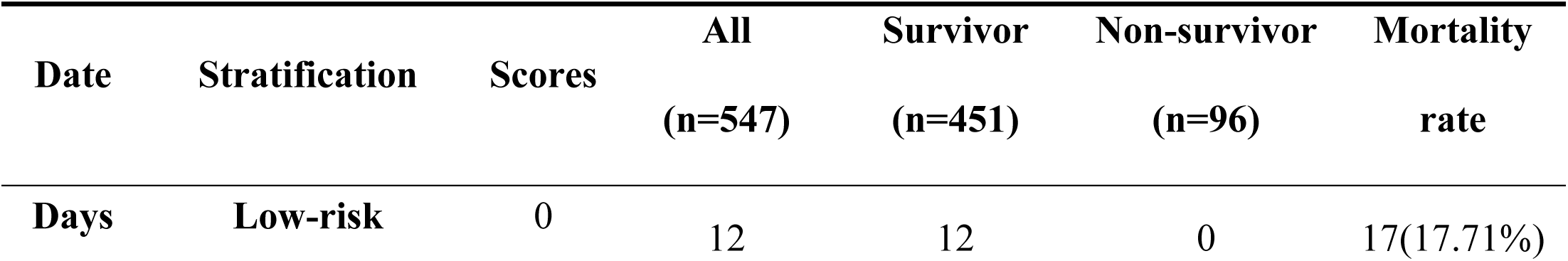
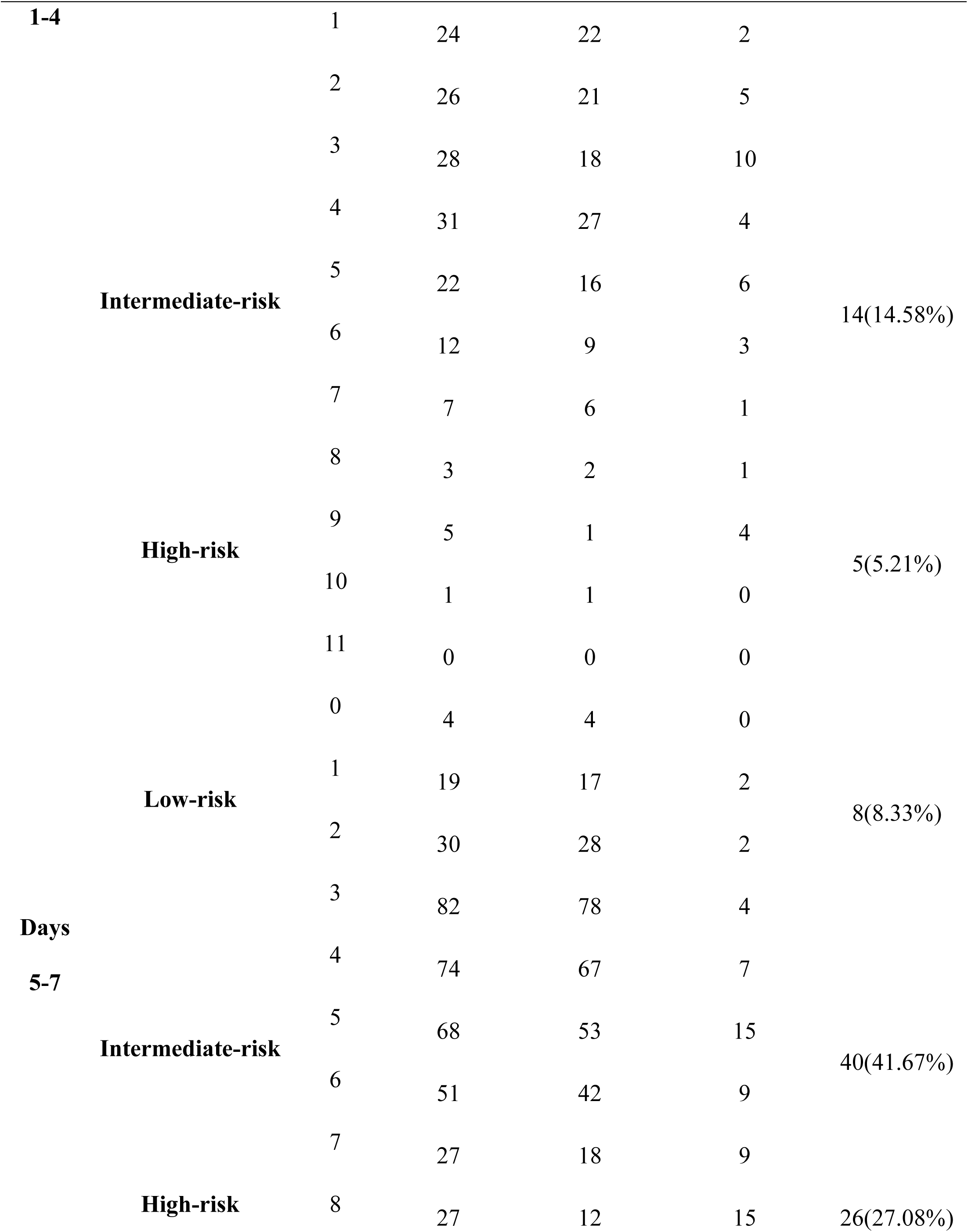
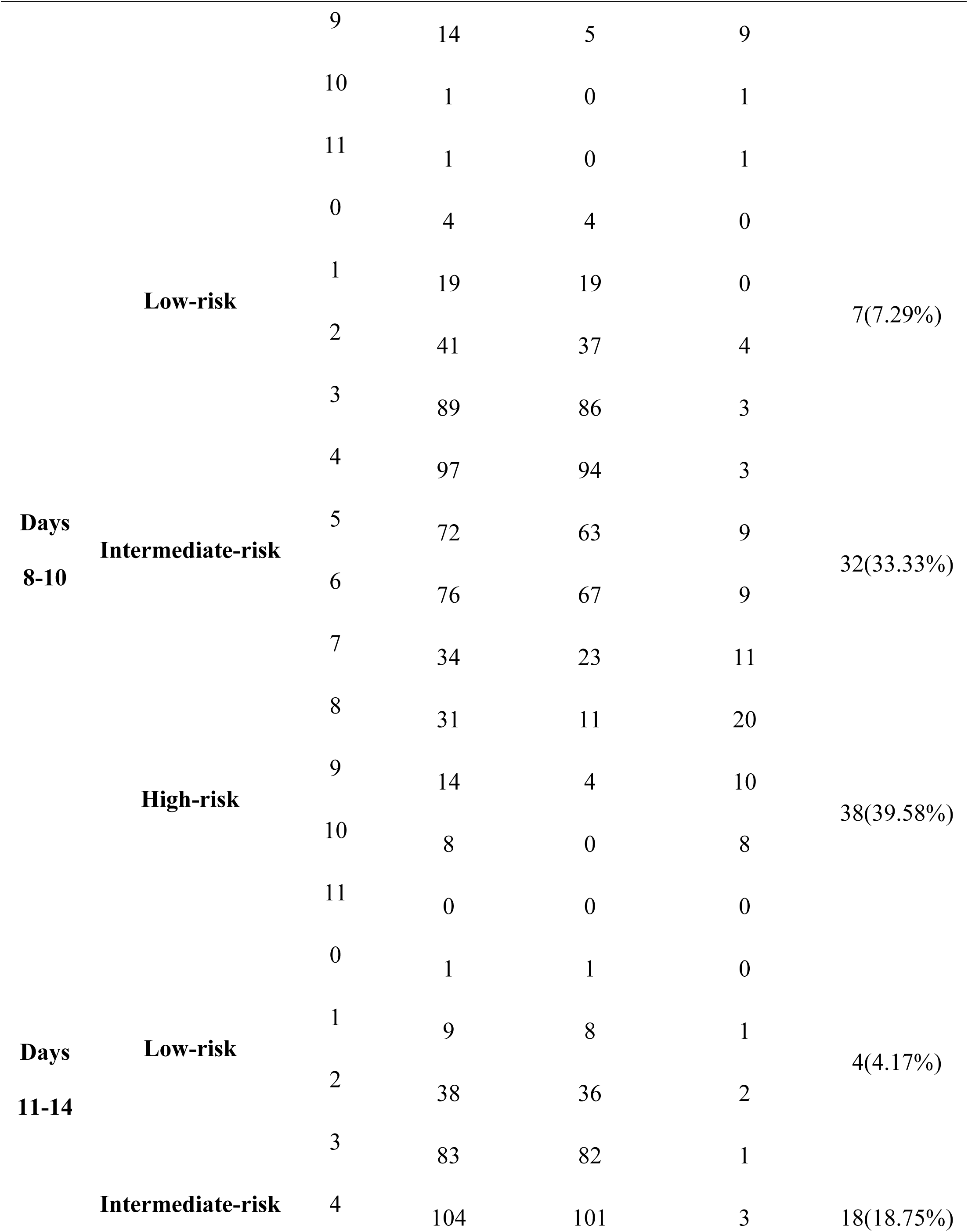
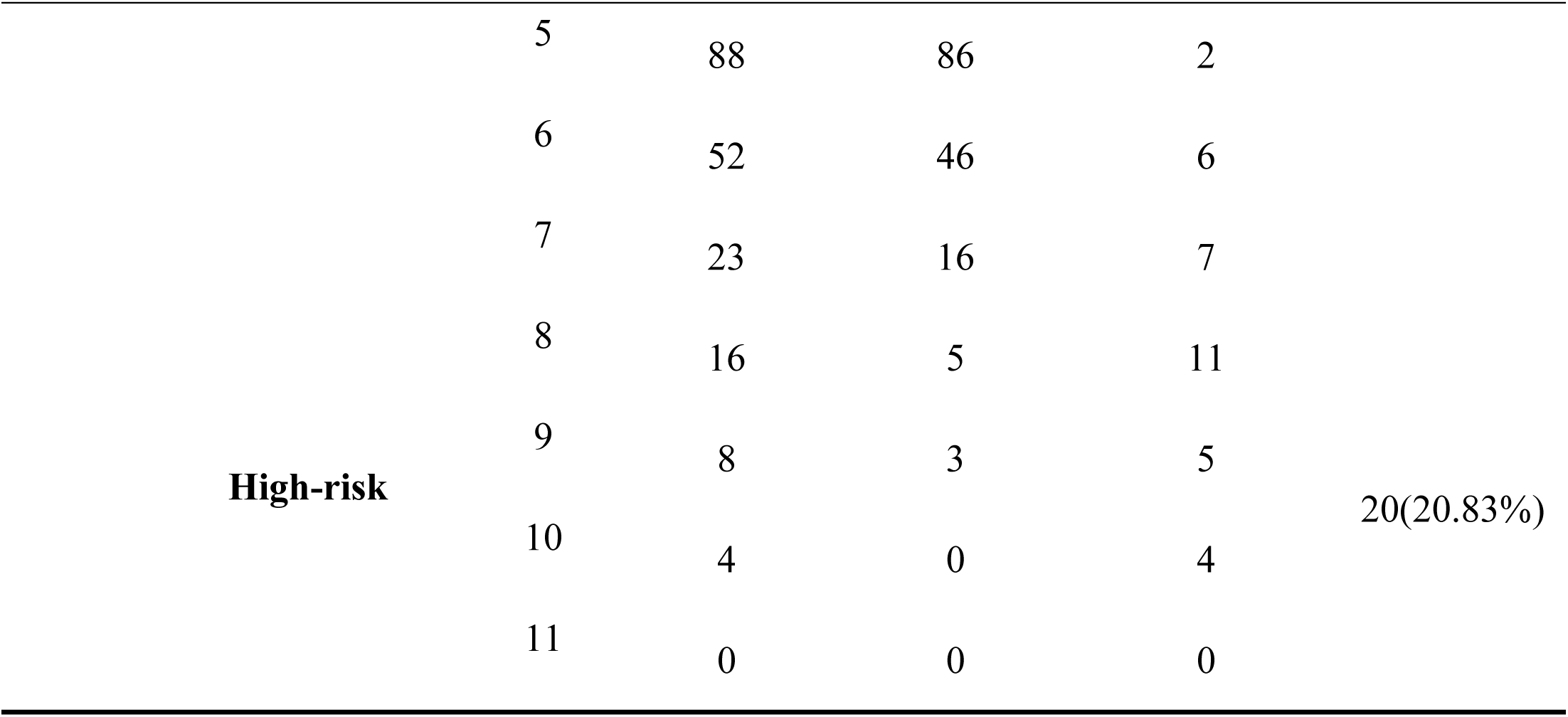
Dynamic Risk Stratification Scores of SFTS Patients During Different.

Days 1-4: Low-risk stratum mortality rate 17.71%, intermediate-risk stratum 14.58%, high-risk stratum 5.21%. The association between risk stratification and mortality at this stage is relatively weak, potentially due to early-stage disease indicators not yet fully manifesting abnormalities.

Days 5-7: Mortality in the intermediate-risk group rose to 41.67%, while the high-risk group reached 27.08%. The low-risk group maintained a low mortality rate of 8.33%, indicating that changes in progressive indicators began to clearly reflect disease severity.

Days 8-10: High-risk group mortality peaked at 39.58%, intermediate-risk group at 33.33%, and low-risk group at only 7.29%. This phase saw concentrated manifestation of multi-organ dysfunction, with risk stratification demonstrating its most pronounced predictive value for prognosis.

Days 11-14: Mortality rates declined across all risk groups, with low-risk at 4.17%, intermediate-risk at 18.75%, and high-risk at 20.83%. This progression aligns with the pathological course of disease improvement and organ function recovery during the remission phase.

### 3.6 Survival Analysis by Severity Stratification

Survival analysis revealed (Table 7) that the overall survival rate in the low-risk group reached 99.00%, with a mean survival time of 31.684 days; the intermediate-risk group had an overall survival rate of 93.10% with a mean survival time of 34.598 days; while the high-risk group recorded an overall survival rate of merely 43.00%, with a mean survival time of 17.505 days and a median survival time of only 8 days. The log-rank test yielded χ²=163.658, degrees of freedom=2, and P<0.001, indicating statistically significant differences in survival curves among the low-, intermediate-, and high-risk groups. This demonstrates the scoring system’s effective risk stratification capability (Fig 2).

**Figure 2.**
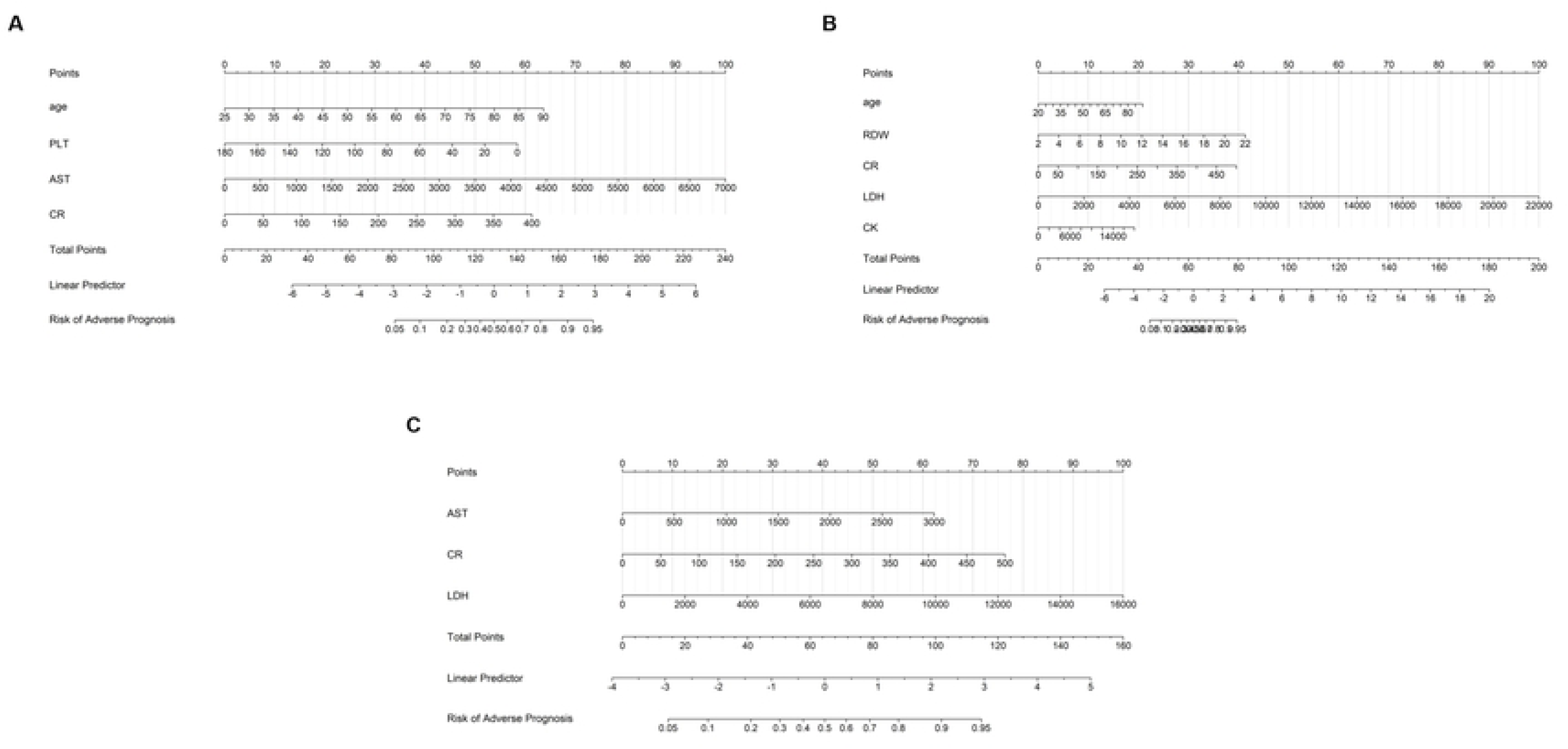
Kaplan-Meier Curves. Kaplan-Meier Curves for overall survival of SFTS patients stratified by the novel severity scoring system. (1)Low-risk; (2)Intermediate-risk; (3)High-risk. Log-Rank test result: χ²=163.211, degrees of freedom=2, P<0.001.

**Table 7.**
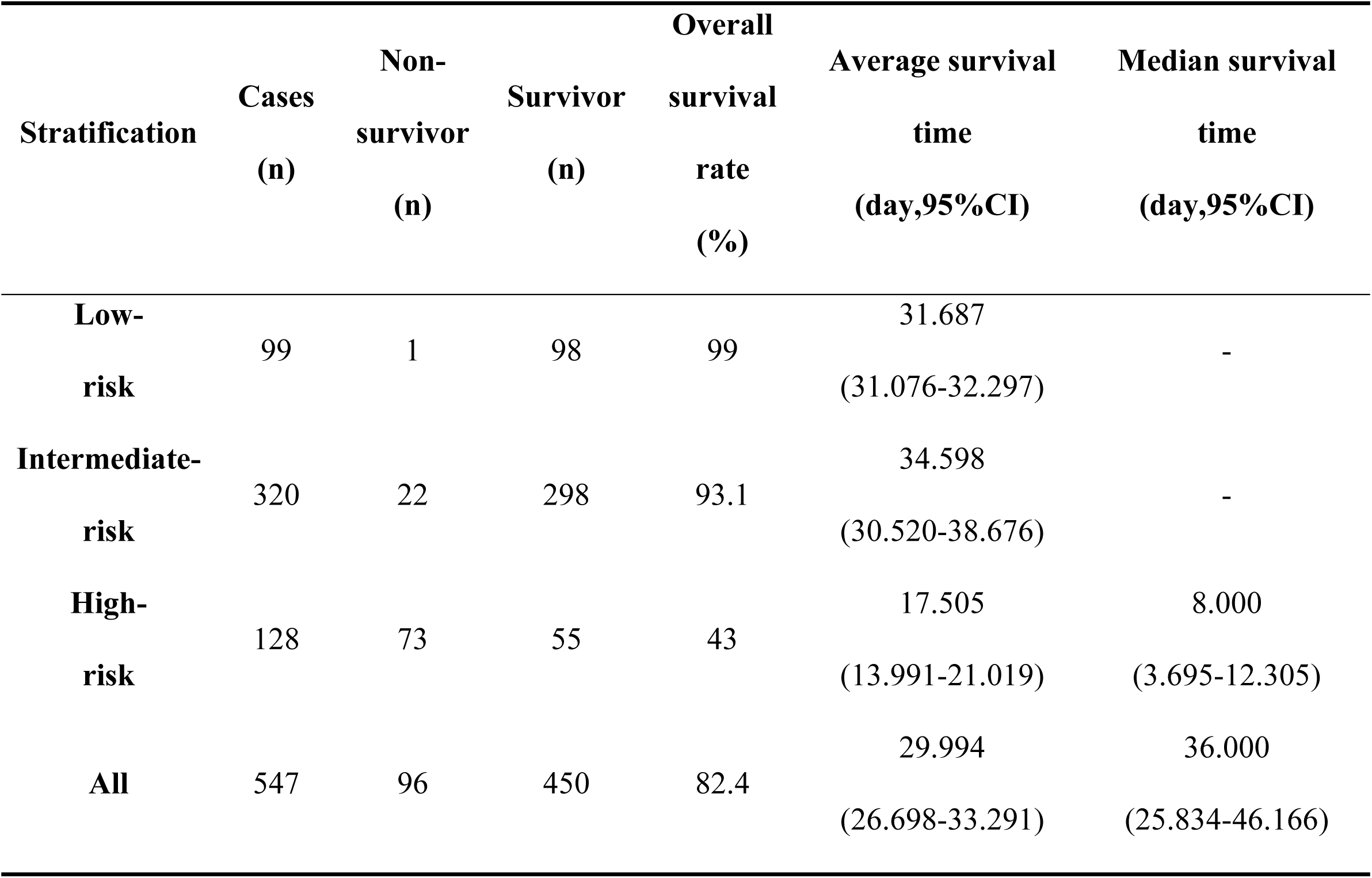
Survival Analysis of SFTS Patients Stratified by Disease Severity. Log-Rank test result: χ ² =163.658, degrees of freedom=2, P<0.001 (indicating statistically significant differences among the three survival curves); “-” indicates that the median survival time was not reached for this group during the follow-up period (i.e., over 50% of patients survived beyond the upper limit of follow-up); 95% CI: 95% confidence interval.

### 3.7 Validation of the Severity Scoring System

Model validation results from the external validation cohort of 44 patients (Table 8) demonstrate the scoring system’s excellent predictive performance:

**Table 8.**
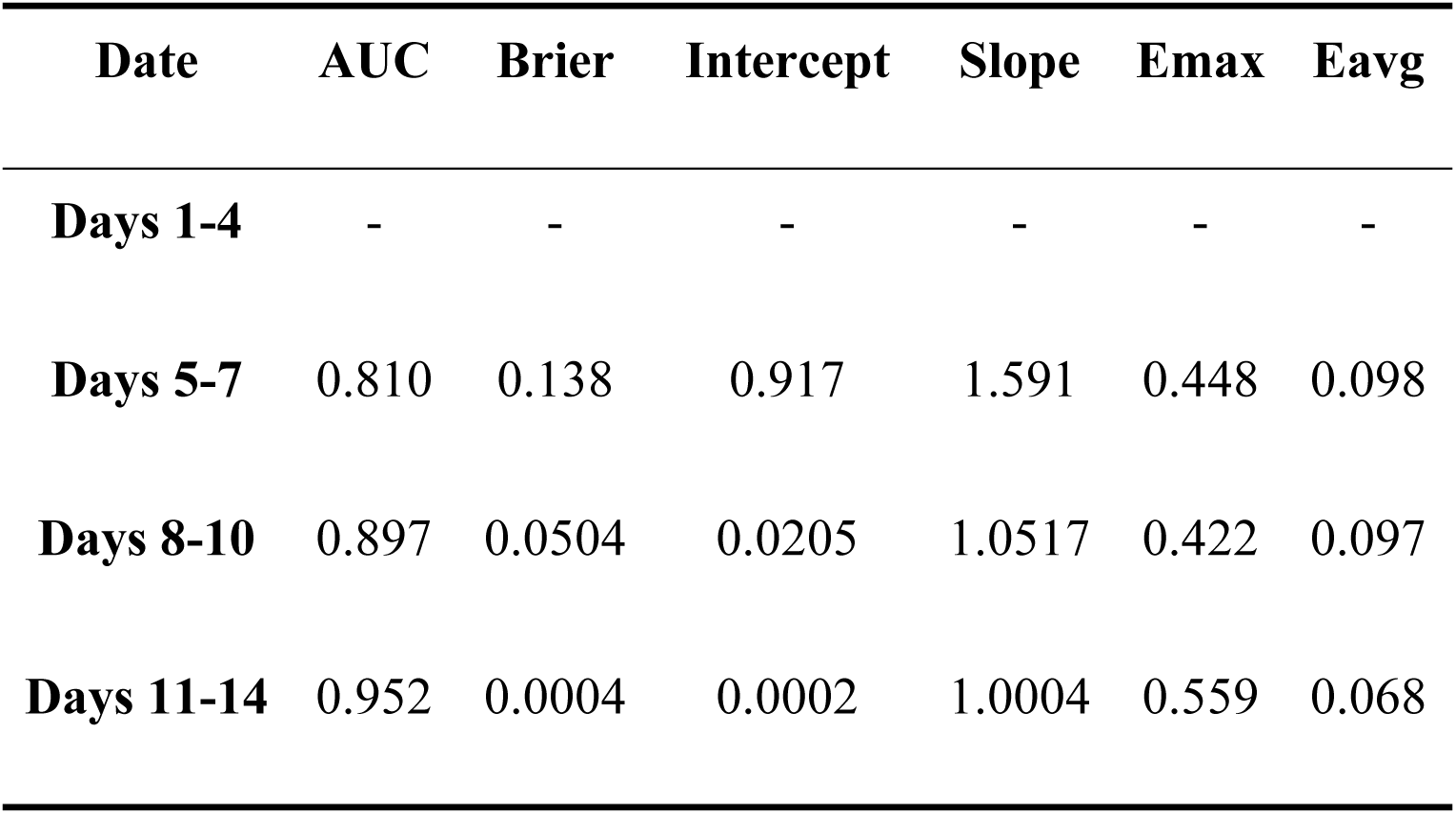
Performance metrics of the SFTS severity scoring system across disease.

Discrimination: AUC values ranged from 0.810 to 0.952 across disease phases, with predictive accuracy progressively improving as the disease advanced. Notably, the 11-14 day phase achieved an AUC of 0.952, indicating optimal model predictive efficacy during this phase (Fig 3).

**Figure 3.**
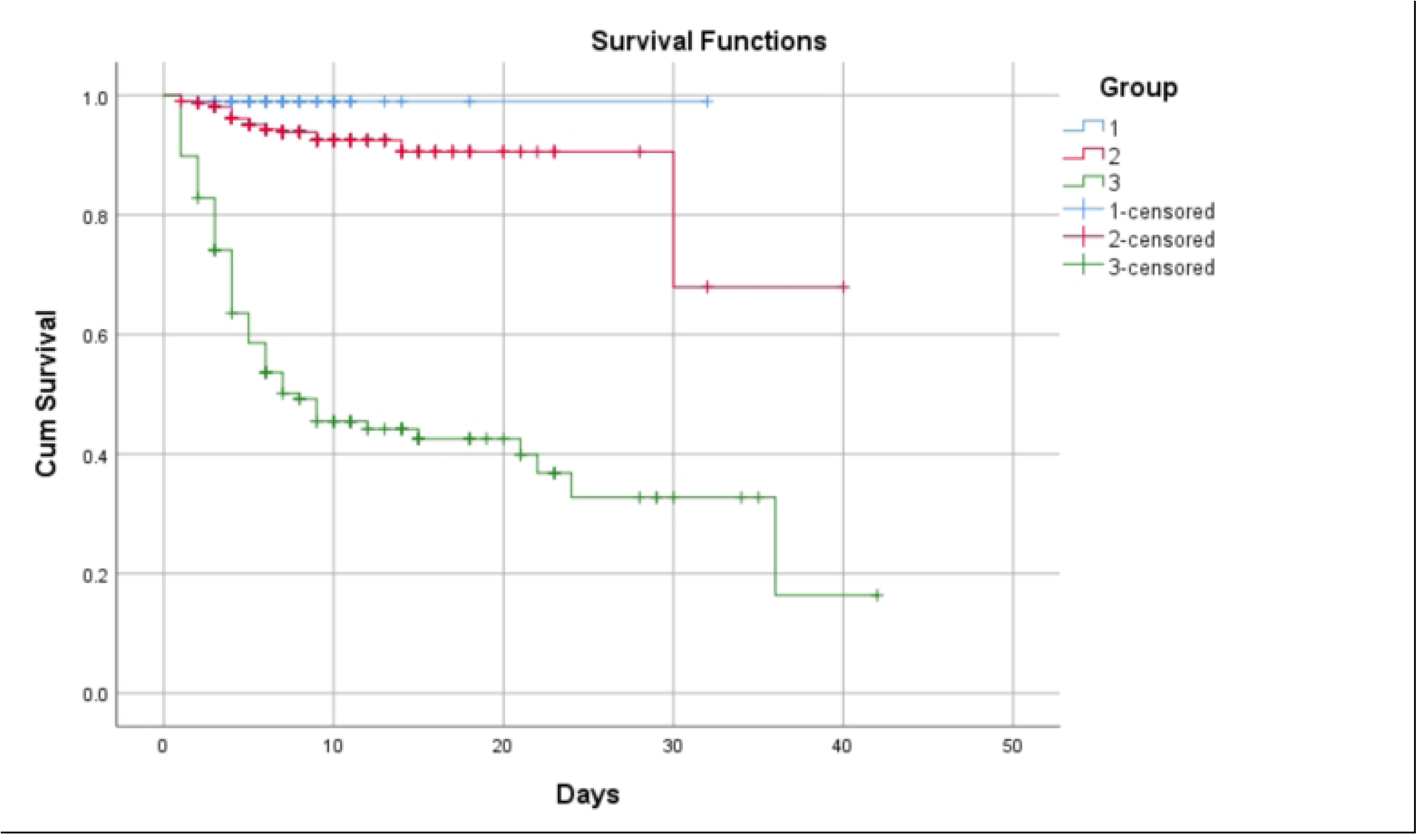
Receiver operating characteristic (ROC) curves. Receiver operating characteristic (ROC) curves for evaluating the discriminatory power of the SFTS severity scoring system across different disease phases. (A)progressive phase; (B)MOD phase; (C)remission phase.

Calibration: The intercept and slope of the calibration curve fell within reasonable ranges, with the Hosmer-Lemeshow test P>0.05, indicating high concordance between predicted probabilities and actual patient outcomes (Fig 4).

**Figure 4.**
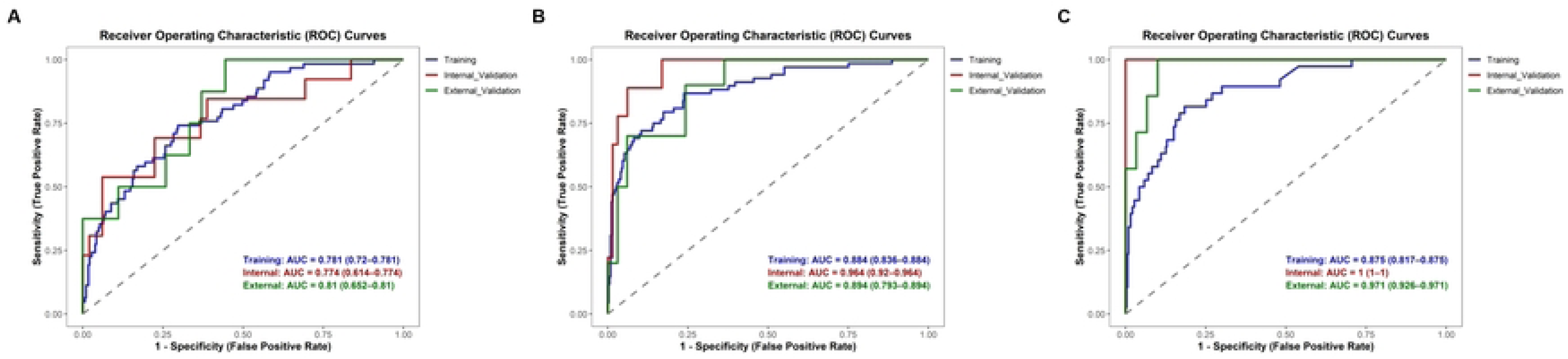
Calibration Curves. Calibration Curves for evaluating the concordance between predicted and actual poor prognosis probabilities of the SFTS severity scoring system across different disease phases. (A)progressive phase; (B)MOD phase; (C)remission phase; (1)Training set; (2)Internal validation set; (3)External validation set.

Clinical net benefit: DCA revealed that DCA curves for all phases exceeded the extreme curve. The mean net benefit (Eavg) ranged from 0.068 to 0.098, with a maximum net benefit (Emax) of 0.422 to 0.559. This effectively mitigates clinical intervention bias, providing reliable support for clinical decision-making (Fig 5).

**Figure 5.**
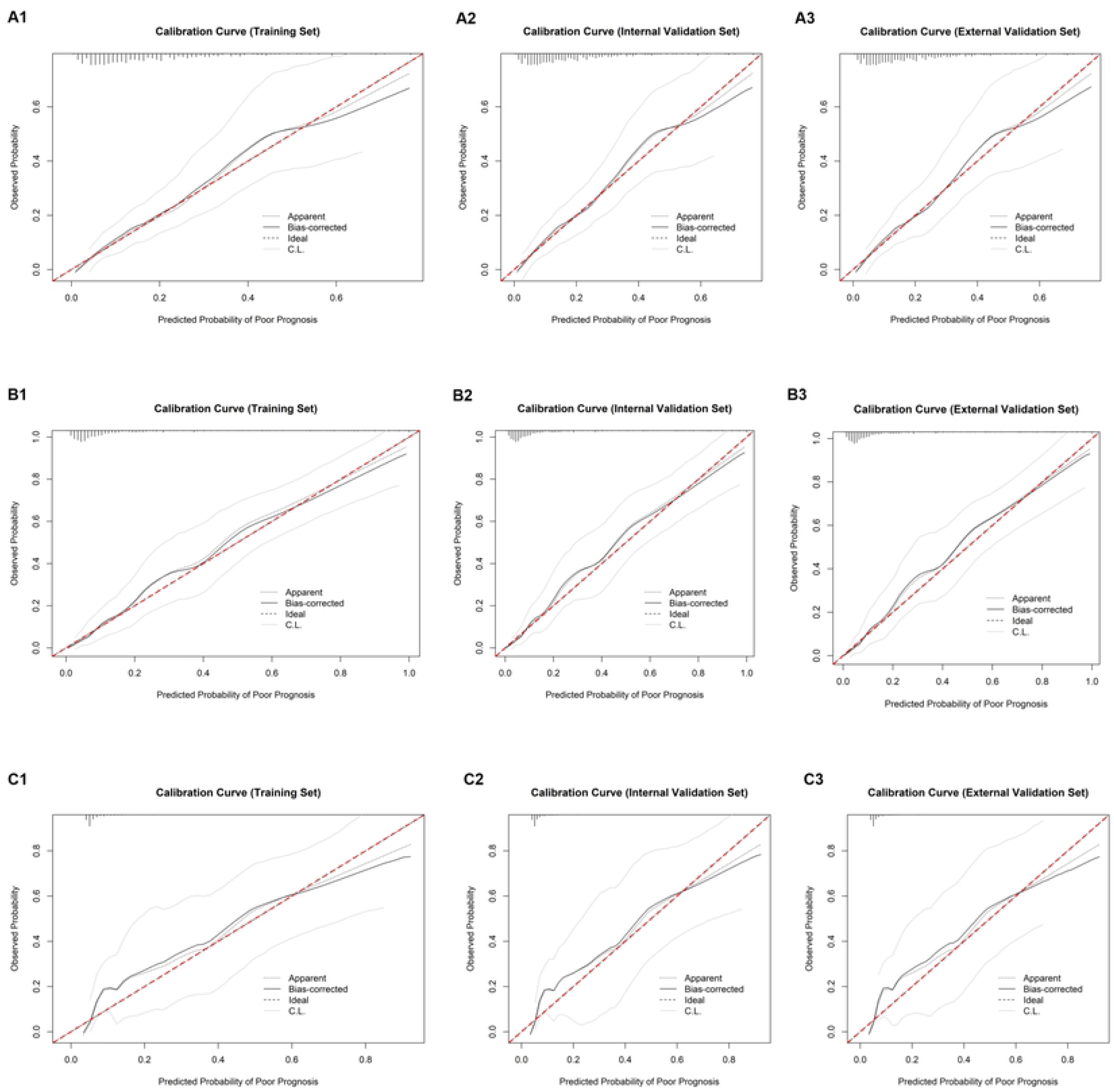
Decision curve analysis (DCA) Decision curve analysis (DCA) for evaluating the clinical net benefit of the SFTS severity scoring system across different disease phases. (A)progressive phase; (B)MOD phase; (C)remission phase.

## Discussion

Severe Fever with Thrombocytopenia Syndrome (SFTS) is an emerging acute infectious disease caused by DBV infection, primarily transmitted by ticks bites but also through contact with bodily fluids and blood of SFTS patients[2,6,25]. It is characterized by high mortality rates and widespread transmission[4,19,20]. The primary clinical symptoms of SFTS include fever, fatigue, digestive system symptoms, and hepatic and renal insufficiency. Severe cases may also present with central nervous system symptoms, disseminated intravascular coagulation (DIC), MOD, and even death[1,3,8,11,25]. Currently, no specific antiviral therapies are available for SFTS, and clinical management primarily relies on symptomatic and supportive care[6,7,26]. Therefore, early identification of the disease severity and the implementation of stratified interventions are crucial for improving patient prognosis[22,23,27].

To date, several studies have developed prognostic assessment tools for SFTS, yet most rely solely on laboratory data at the single time point of hospital admission. These tools typically categorize results as normal or abnormal based solely on reference ranges, overlooking the dynamic progression of the disease[22,24]. Consequently, they fail to provide effective guidance for adjusting intervention strategies during the course of illness[8,9,10,23,27]. For instance, some patients admitted in the early disease phase may exhibit no typical abnormalities in relevant indicators, leading to misclassification as low-risk. Conversely, others experiencing rapid indicator changes during the mid-disease phase may suffer treatment delays due to the absence of dynamic monitoring[11,21,22].

This study divided the 14-day disease course into four key phases, extracting the extreme values of laboratory findings at each phase (such as the lowest platelet count and peak AST levels) to more precisely capture the patterns of disease progression[8,9,24,26,27]. The pathophysiological process of SFTS is dynamically evolving: the initial phase is characterized by systemic inflammatory responses triggered by viral replication; the intermediate phase involves progressive platelet destruction and organ dysfunction (such as hepatic and renal insufficiency); the late phase may result in fatal outcomes due to severe inflammatory storms or multiple organ failure[6,14,15,16,19,20,25]. Findings measured at a single time point (e.g., upon admission) reflect only the disease state at that specific phase. This approach may yield inaccurate assessments if patients exhibit no typical abnormalities during the early disease phase or if indicators temporarily decline during compensatory periods[21,22,23,24].

Through LASSO regression and multivariate logistic regression analysis, we dynamically identified six independent prognostic risk factors—age, RDW, PLT, AST, Cr, and LDH—with distinct distributions across disease phases[8,9,10,24]. Elderly patients (age>67 years) exhibit compromised immune function and diminished viral clearance capacity, rendering them more susceptible to severe disease progression[4,5,11,13,19]. Reduced PLT counts indicate significant excessive platelet consumption; prior studies suggest this may relate to transient suppression of bone marrow hematopoiesis induced by viral infection. Additionally, the binding of PLT to SFTSV further promotes their clearance by splenic macrophages, thereby increasing the patient’s risk of hemorrhage and MODS[8,9,10,13,14]. Elevated AST and LDH reflect hepatocyte and cardiomyocyte injury, serving as biomarkers of worsening hepatic damage and systemic inflammatory response[5,8,17,18,22]. Increased Cr levels are indicative of renal impairment, a critical hallmark of multi-organ failure in severe SFTS[14,15]. Elevated RDW suggests abnormal erythropoiesis, which correlates with systemic inflammation and poor prognosis, potentially reflecting the indirect effects of viral-induced inflammation on bone marrow function[12,15,16,24].

A key innovation of this study lies in its refined scoring methodology. Unlike conventional scoring systems that adopt arbitrary equal-point allocation without accounting for the clinical relevance of indicator ranges, we utilized the Youden index from receiver operating characteristic (ROC) curves to determine evidence-based clinical cut-off values for each risk factor[9,22,23,24]. Integrating these cut-offs with clinically validated normal reference ranges, we developed a stratified scoring rule that links point weights to the severity of clinical risk. For example, a PLT count <50 ×10⁹/L (high risk) was assigned 2 points, 50-100×10⁹/L (medium risk) 1 point, and 100-300×10⁹/L (low risk) 0 points. This approach ensures that the scoring system is both scientifically rigorous and clinically meaningful, avoiding over– or under-weighting of indicators based on arbitrary criteria.

To enhance clinical applicability, we further stratified total scores (0-11 points) into low-risk (0-3 points), intermediate-risk (4-7 points), and high-risk (8-11 points) tiers based on quartile analysis. Primary healthcare institutions can rapidly complete clinical assessments in three steps: assigning scores to indicators by tier, summing total scores, and performing risk stratification. This enables early evaluation of SFTS patients’ conditions. Unlike previous complex prognostic models (such as multivariate nomograms)[21,22,23,27], this scoring system is more concise, straightforward and practical, addressing the unmet need for simple tools in primary care settings[6,20]. It provides a useful tool for stratifying SFTS severity and facilitating timely intervention.

The reliability and clinical utility of the scoring system were validated through a comprehensive multidimensional evaluation. In the development cohort, K-M curves and log-rank tests demonstrated statistically significant differences in mortality across risk tiers (log-rank P<0.001), confirming the system’s ability to distinguish between patients with varying disease severity. In the external validation cohort, the C-statistic, calibration curves, and decision curve analysis (DCA) collectively validated the systems robust discriminatory power (AUC=0.810-0.952), excellent calibration (Hosmer-Lemeshow test P>0.05), and substantial clinical net benefit (Eavg=0.068-0.098, Emax=0.422-0.559). Importantly, the DCA curves consistently outperformed extreme strategies (treating all or no patients as high-risk), effectively mitigating the risks of over-intervention (unnecessary intensive care for low-risk patients) and under-intervention (delayed treatment for high-risk patients) in clinical decision-making[6,21,23,27].

Compared with existing SFTS scoring tools, our dynamic scoring system offers several key advantages. By incorporating phase-specific extreme values and evidence-based cut-offs, it more accurately reflects the dynamic progression of SFTS, enabling timely adjustments to intervention strategies[22,23,24]. This has significant implications for clinical practice: Low-risk patients may undergo outpatient follow-up and home isolation, minimizing unnecessary healthcare resource utilization and avoiding overtreatment[6,27]. Intermediate-risk patients require close monitoring of dynamic changes in indicators (such as daily re-evaluation of PLT, hepatic function and renal function), with timely adjustments to treatment regimens and symptomatic management[8,9,10,17]. High-risk patients necessitate early initiation of intensive care, including immunomodulatory therapy, supportive treatment (such as platelet transfusion), and hepatoprotective and nephroprotective interventions, to reduce the risk of MODS and improve survival[1,4,11,13].

Notably, our findings highlight that the MOD phase (days 8-10) is the most critical period for prognosis, with the highest number of independent risk factors and peak mortality rates[9,11,24]. This underscores the importance of intensified monitoring and intervention during this phase, as timely organ support may prevent irreversible multi-organ damage[4,13]. Additionally, the identification of Cr and LDH as persistent independent risk factors throughout the progressive, MOD, and remission phases emphasizes the central role of renal and myocardial injury in SFTS prognosis, providing targets for focused organ protection[5,6,8,10,19].

This study also has certain limitations: Firstly, missing data were imputed using median imputation, which, although reducing data loss, may introduce some error; future studies could reduce data loss rates by improving clinical record-keeping. Secondly, the study did not include potential confounding factors (such as antiviral drug use); subsequent research could expand the range of variables(e.g., treatment regimens, troponin levels, viral load) to further optimize the scoring system. Furthermore, the external validation cohort comprised a relatively small number of cases. Future studies should incorporate multicenter data for further external validation to enhance the scoring system’s generalizability.

## Conclusion

Age, RDW, AST, LDH, Cr and PLT constitute independent risk factors for prognosis in SFTS patients. This study developed and validated a dynamic, phase-specific scoring system for SFTS severity quantification, based on 14-day clinical data and evidence-based cut-offs. The system effectively stratifies patients into low-, intermediate-, and high-risk tiers, with robust reliability and substantial clinical utility. By enabling early, precise risk assessment and individualized intervention, this tool addresses an unmet clinical need in SFTS management, potentially improving patient prognosis and optimizing healthcare resource allocation.

## Author Statement

Yingchun Sun: Conceptualization, Formal Analysis and Writing-Original Draft.

Jing Sun and Zhiyu Pan: Design, Data Collection and Writing-Original Draft.

Yihui Sun, Qiongle Wu and Haoyu Sheng: Writing-Review & Editing.

Wenjie Wang, Manman Liang, Aiping Zhang: Data Collection.

Jianghua Yang: Conceptualization, Methodology and Writing-Review & Editing.

All authors commented on previous versions of manuscript. All authors read and approved the final manuscript.

## Data Availability

The data that support the findings of this study are available from the corresponding author upon reasonable request.

## Acknowledgements

We would like to express our gratitude to all patients and medical staff who participated in this study.

## Ethics Statement

The study was conducted according to the guidelines of the Declaration of Helsinki. A statement to confirm that all experimental protocols were approved by the Ethics Committee of Scientific Research and New Technology of Wannan Medical College Yijishan Hospital Institutional Review Board (IRB; Research Proposal Notification IRB Review Decision No. (2023) 05, dated June 20, 2023. The requirement for informed consent was waived by the Ethics Committee of Scientific Research and New Technology of Wannan Medical College Yijishan Hospital IRB because of the retrospective nature of the study.

## References

[1] Seo JW, Kim D, Yun N, Kim DM. Clinical Update of Severe Fever with Thrombocytopenia Syndrome. Viruses. 2021;13(7):1213. Published 2021 Jun 23. doi:10.3390/v13071213

[2] Sharma D, Kamthania M. A new emerging pandemic of severe fever with thrombocytopenia syndrome (SFTS). Virusdisease. 2021;32(2):220–227. doi:10.1007/s13337-021-00656-9

[3] Luo N, Li M, Xu M, et al. Research Progress of Fever with Thrombocytopenia Syndrome. Intensive Care Res. Published online March 23, 2023. doi:10.1007/s44231-023-00035-6

[4] Dualis H, Zefong AC, Joo LK, et al. Factors and outcomes in Severe Fever with Thrombocytopenia Syndrome (SFTS): A systematic review. Ann Med Surg (Lond). 2021;67:102501. Published 2021 Jun 11. doi:10.1016/j.amsu.2021.102501

[5] Hao Y, Wang X, Du Z, et al. Prevalence and impact of viral myocarditis in patients with severe fever with thrombocytopenia syndrome. J Med Virol. 2024;96(4):e29612. doi:10.1002/jmv.29612.

[6] Peng C, Hao Y, Yuan Y, et al. Bandavirus dabieense: A review of epidemiology, clinical characteristics, pathophysiology, treatment and prevention. Virulence. 2025;16(1):2520343. doi:10.1080/21505594.2025.2520343

[7] National Health and Family Planning Commission of the People’s Replublic China. Guide for severe fever with thrombocytopenia syndrome control. 2010. (edit). [cited 2013 Sept 23].

[8] Wang Y, Song Z, Wei X, et al. Clinical laboratory parameters and fatality of Severe fever with thrombocytopenia syndrome patients: A systematic review and meta-analysis. PLoS Negl Trop Dis. 2022;16(6):e0010489. Published 2022 Jun 17. doi:10.1371/journal.pntd.0010489

[9] He Z, Wang B, Li Y, Du Y, Ma H, Li X, et al. Severe fever with thrombocytopenia syndrome: a systematic review and meta-analysis of epidemiology, clinical signs, routine laboratory diagnosis, risk factors, and outcomes. BMC Infect Dis. 2020; 20(1): 575. doi: 10.1186/s12879-020-05303-0

[10] Li MM, Lei XY, Yu XJ. Meta-analysis of the clinical and laboratory parameters of SFTS patients in China. Virol J. 2016; 13(1): 198. doi: 10.1186/s12985-016-0661-9

[11] Wang X, Ren X, Ge Z, Cui S, Wang L, Chen Z, et al. Clinical manifestations of death with severe fever and thrombocytopenia syndrome: A meta-analysis and systematic review. J Med Virol. 2021; 93(6): 3960–3968. doi: 10.1002/jmv.26518

[12] Dualis H, Zefong AC, Joo LK, Dadar Singh NK, Syed Abdul Rahim SS, Avoi R, et al. Factors and outcomes in Severe Fever with Thrombocytopenia Syndrome (SFTS): A systematic review. Ann Med Surg (Lond). 2021; 67: 102501. doi: 10.1016/j.amsu.2021.102501

[13] Chen Y, Jia B, Liu Y, Huang R, Chen J, Wu C. Risk factors associated with fatality of severe fever with thrombocytopenia syndrome: a meta-analysis. Oncotarget. 2017;8(51):89119–89129. Published 2017 Jul 11. doi:10.18632/oncotarget.19163

[14] Jin C, Liang M, Ning J, Gu W, Jiang H, Wu W, et al. Pathogenesis of emerging severe fever with thrombocytopenia syndrome virus in C57/BL6 mouse model. Proc Natl Acad Sci USA. 2012; 109(25): 10053–10058. doi: 10.1073/pnas.1120246109

[15] Yang M, Zhang Y, Zhang Y, et al. Investigation of clinical characteristics and prognostic factors of severe fever with thrombocytopenia syndrome: an analysis of 69 cases. J Dis Control Prev. 2018;22(4):350–354. doi:10.16462/j.cnki.zhjbkz.2018.04.018.

[16] Yu XJ. Risk factors for death in severe fever with thrombocytopenia syndrome. Lancet Infect Dis. 2018; 18(10): 1056–1057. doi: 10.1016/S1473-3099(18)30312-8

[17] Wang L, Liu XZ, Lin Q. Prognostic value of laboratory parameters in patients with severe fever thrombocytopenia syndrome. Inter J Epidemiol Infect Dis. 2017; 44(6):370–373. doi: 10.3760/cma.j.issn.1673-4149.2017.06.003

[18] Shao F, Zhang Y, Hu XM, Zhang LH, Bian PF, Li Q. Characteristics and significances of ECG in patients of severe fever with thrombocytopenia syndrome. Chin J Exp Clin Infect Dis (Electron Ed). 2015;9(6):763–766.

[19] Li H, Lu QB, Xing B, Zhang SF, Liu K, Du J, et al. Epidemiological and clinical features of laboratory-diagnosed severe fever with thrombocytopenia syndrome in China, 2011–2017: a prospective observational study. Lancet Infect Dis. 2018; 18(10):1127–1137. doi: 10.1016/S1473-3099(18)30293-7

[20] Sun H, Hu Q, Lu S, et al. Current status of severe fever with thrombocytopenia syndrome in China (Review). Int J Mol Med. 2025;56(5):169. doi:10.3892/ijmm.2025.5610

[21] Wang L, Xie H, Liu Y, Zou Z. A nomogram including admission serum glycated albumin/albumin ratio to predict mortality in patients with severe fever with thrombocytopenia syndrome. BMC Infect Dis. 2024;24(1):858. Published 2024 Aug 23. doi:10.1186/s12879-024-09752-9

[22] Zhang Y, Zhong P, Wang L, et al. Development and validation of a clinical risk score to predict the occurrence of critical illness in hospitalized patients with SFTS. J Infect Public Health. 2023;16(3):393–398. doi:10.1016/j.jiph.2023.01.007

[23] Zhong F, Lin X, Zheng C, et al. Establishment and validation of a clinical risk scoring model to predict fatal risk in SFTS hospitalized patients. BMC Infect Dis. 2024;24(1):975. Published 2024 Sep 13. doi:10.1186/s12879-024-09898-6

[24] Liu Z, Jiang Z, Zhang L, et al. A model based on meta-analysis to evaluate poor prognosis of patients with severe fever with thrombocytopenia syndrome. Front Microbiol. 2024;14:1307960. Published 2024 Jan 8. doi:10.3389/fmicb.2023.1307960

[25] Liu Q, He B, Huang SY, Wei F, Zhu XQ. Severe fever with thrombocytopenia syndrome, an emerging tick-borne zoonosis. Lancet Infect Dis. 2014;14(8):763–772. doi:10.1016/S1473-3099(14)70718-2.

[26] Li JC, Zhao J, Li H, Fang LQ, Liu W. Epidemiology, clinical characteristics, and treatment of severe fever with thrombocytopenia syndrome. Infect Med (Beijing). 2022;1(1):40–49. Published 2022 Jan 1. doi:10.1016/j.imj.2021.10.001.

[27] Chen G, Du Y, Ma X, et al. Retrospective Analysis of Severe Fever With Thrombocytopenia Syndrome and Construction of a Nomogram Prediction Model for Mortality Risk Factors. Open Forum Infect Dis. 2025;12(7):ofaf318. Published 2025 Jun 2. doi:10.1093/ofid/ofaf318

